# The association between body roundness index and kidney stones in a diabetic population: NHANES 2011–2018

**DOI:** 10.1101/2024.12.12.24318931

**Authors:** Chongsong Cui, You Peng, Ying Zhao, Feiin Chan, Qiqi Ren, Zhenjie Liu

## Abstract

**Background:** Predominantly attributed to metabolic disruptions, individuals with diabetes are more prone to kidney stones than the general population. The Body Rounds Index (BRI), a new measure of obesity and health risks, has been shown to have a positive correlation with the risk of developing kidney stones. Our study sets out to explore the correlation between BRI and kidney stones specifically within the diabetic population.

**Methods:** Leveraging data from the National Health and Nutrition Examination Survey (NHANES) spanning 2011 to 2018, this cross-sectional study probed the link between BRI and kidney stones among diabetic individuals. We conducted a logistic regression analysis to investigate the relationship between BRI and kidney stones, with adjustments for various covariates. To further explore the association trends between different BRI levels and the incidence of kidney stones, we categorized BRI into four levels, thereby enhancing the robustness of our results. Additionally, subgroup and sensitivity analyses were performed to ensure the reliability and consistency of our findings.

**Results:** A total of 3,558 diabetic patients were included in this study, of whom 546 (15.3%) had kidney stones. After adjusting for some and all confounders, we observed that the prevalence of kidney stones in diabetic patients increased by 7% and 5% for each unit increase in BRI in Models 2 and 3, respectively (Model 2: OR=1.07, 95% CI: 1.03-1.11; Model 3: OR=1.05, 95% CI: 1.00-1.09).This association remained statistically significant when the BRI was divided into quartiles, the prevalence of kidney stones in the highest quartile of BRI (Q4) was 50% greater than in the lowest quartile (Q1) (OR=1.50, 95% CI: 1.14-1.98; P<0.05).

**Conclusions:** Our research indicates a positive correlation between BRI and the incidence of kidney stones among diabetics. We believe that BRI may be an effective indicator for assessing the risk of kidney stones in diabetic patients.

## 1. Introduction

Kidney stones, a common urological disorder, are solid crystalline deposits that form due to the abnormal accumulation of minerals and acid salts in the kidneys(1). The incidence of kidney stones worldwide stands at around 10% and is persistently climbing(1,2). Extracorporeal shock wave lithotripsy and other minimally invasive procedures are effective methods for stone extraction; however, the recurrence rate of kidney stones can be as high as 50%(3). Kidney stones can lead to serious complications, including urinary tract infections, kidney damage, and even renal failure(4), which present significant challenges to public health systems. Thus, identifying risk factors for kidney stones and implementing effective preventive measures is crucial(2,5).

Studies have shown that the incidence of kidney stones in the diabetic population is significantly higher than that in the general population(6), which aroused our concern, which may be attributed to many factors such as disorders of glucose-fat metabolism and obesity, and Chou YH et al. also proved that there is a positive correlation between the incidence of body weight gain and KSD(7), and as the mainstream indicator for assessing obesity——Body Mass Index(BMI), current research has confirmed that BMI does not accurately reflect the distribution of body fat (8).Consequently, Thomas et al. proposed the Body Roundness Index (BRI) as an emerging measure for evaluating obesity and associated health risks. BRI is calculated by combining height and waist circumference, allowing for a more effective assessment of visceral fat tissue and overall body fat percentage(9). Research by Xudong Hu et al. (10)found a linear positive correlation between BRI and the risk of developing kidney stones, with BRI showing a significantly greater ability to differentiate kidney stone risk than BMI and waist circumference. Xike Mao et al. (11)established that for each unit increase in BRI, the prevalence of kidney stones increased by 65% (OR = 1.65, 95% CI: 1.47, 1.85) after adjusting for confounding factors. However, most of these studies have focused on the general population, and research examining the relationship between BRI and the prevalence of kidney stones in specific populations, such as diabetic patients, remains relatively limited. Research by Liting Qiu et al. (12)showed that patients with diabetes and prediabetes had higher BRI values (Mean ± SD = 5.89 ± 2.21) than the non-diabetic population (Mean ± SD = 4.63 ± 1.85), and the elevated BRI values further affected their risk of renal stones. Therefore, by identifying and intervening on BRI, it can help to more accurately assess the risk of kidney stones in diabetic patients and may lead to more effective personalized prevention and treatment strategies. By monitoring and improving BRI, the goal of reducing the prevalence of kidney stones can be achieved, especially in the high-risk population of diabetes.

Although current studies have demonstrated an association between BRI and kidney stones, these studies have focused on the general population, and there is a lack of in-depth research on the relationship between BRI and kidney stones in diabetic patients. Because the association between BRI and kidney stones in the diabetic population is unknown, we sought to reveal the specific role of BRI in the development of diabetes-associated kidney stones and to provide new strategies and perspectives for the prevention and treatment of kidney stones through a cross-sectional study of diabetic patients based on the NHANES survey conducted in the United States between 2011 and 2018.

## 2. Methods

### 2.1 Study participants

The data for this research are based on the NHANES survey conducted between 2011 and 2018. NHANES is a national cross-sectional study conducted in the United States that employs a stratified multistage random sampling methodology to collect data from the ambulatory population. The detailed design, methodology, and data of the survey are available on its official website (http://www.example.com) and the U.S. Centers for Disease Control and Prevention’s (CDC) National Health and Nutrition Examination Survey (NHANES) page (http://www.cdc.gov/nchs/nhanes.htm). All participants provided written consent to participate in the NHANES study, which was approved by the ethics committee(13). Consequently, no additional approvals or ethical reviews were required for this study. Of the initial 4,432 diabetic patients collected, we excluded 74 individuals younger than 20 years of age, 496 individuals with incomplete BRI data, 13 individuals with incomplete information regarding renal stones (including diagnosis), and 291 individuals with missing covariate data. This resulted in a final sample size of 3,558 individuals included in the study (Figure 1).

**Figure 1.**
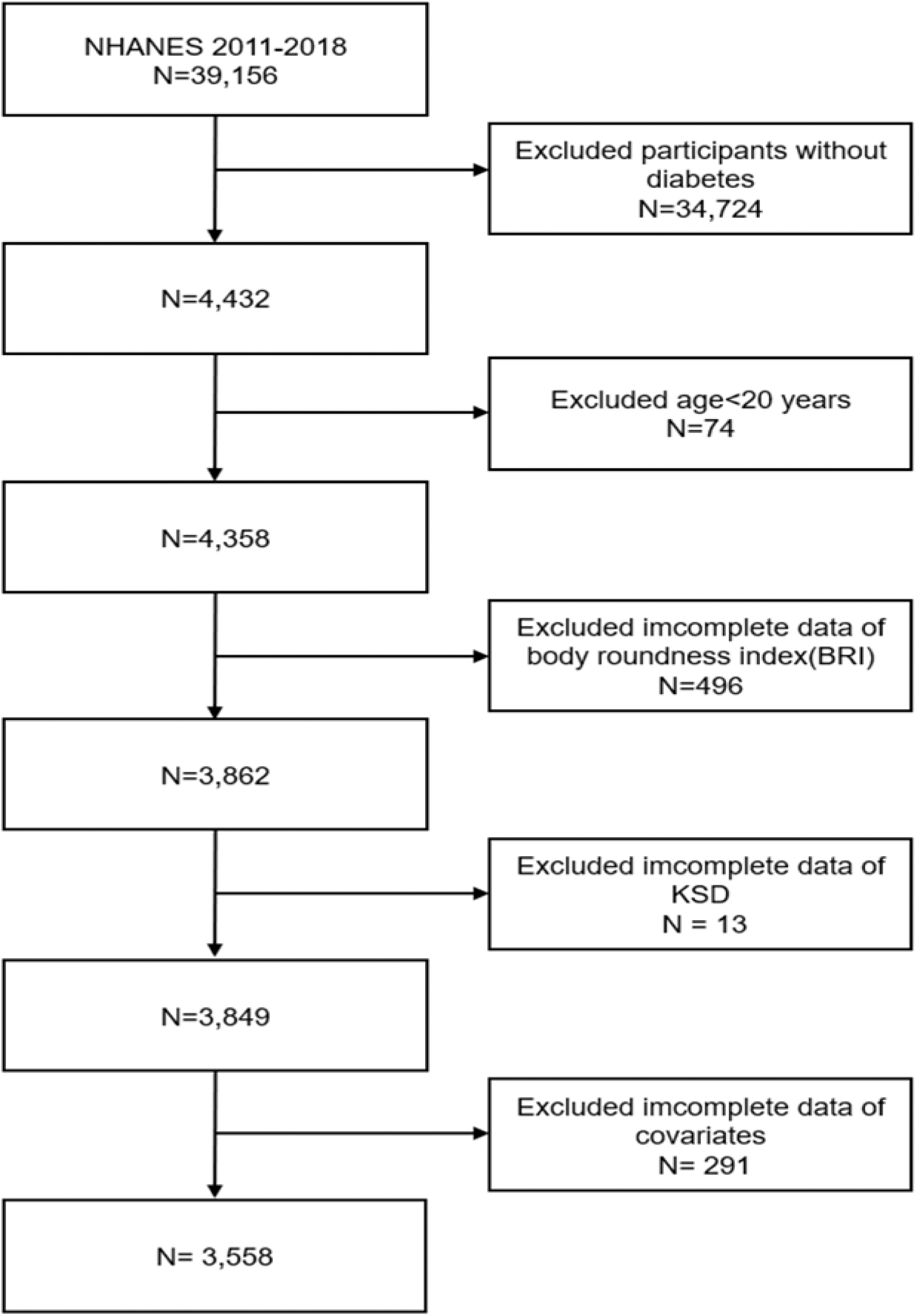
Flow chart of participant selection.

### 2.2 Assessment of body roundness index

In 2013, Thomas and colleagues initially introduced the concept of BRI(9).BRI uses height and waist circumference (WC) to assess the percentage of visceral and total body fat. We obtained participants’ waist circumference (WC) and height data from the body measurements section of the NHANES database. The following formula was then applied to calculate the BRI values:

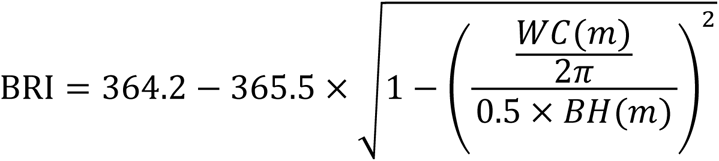

### 2.3 Diabetes and kidney stones

Diabetes is diagnosed in an individual who fulfills one or more of the subsequent criteria: (1) A fasting blood glucose value of at least 7.0 mmol/L or a 2-hour oral glucose tolerance test result of ≥11.1 mmol/L; (2) a blood glucose concentration of ≥11.1 mmol/L at any given time; (3) a glycosylated hemoglobin (HbA1c) level of ≥6.5%; (4) ongoing use of glucose-lowering medications or insulin injections; and (5) a self-reported diabetes diagnosis by a physician(14). The urology section of NHANES includes a questionnaire called the KIQ26, which collects information about participants’ history of kidney stones. Participants’ history was assessed by asking, “Have you ever had a kidney stone?” Participants who responded affirmatively were identified as having a history of kidney stones, while those who answered negatively were categorized as having no such history.

### 2.4 Covariates

Kidney stones are influenced by many factors, we included the following covariates: age, gender, education level (less than 9th grade, 9th-11th grade, high school graduate/GED or equivalent, some college or AA degree, college graduate or above), marital status (married/living with partner, widowed/divorced/separated, never married), smoking status (defined as having smoked at least 100 cigarettes), activity status (vigorous work activity, moderate work activity, vigorous recreational activities, moderate recreational activities, physical activity—walking or bicycling, minutes of sedentary activity), height, weight, and body mass index (BMI), calculated as weight (kg) divided by height (m) squared. Total cholesterol (TC), high-density lipoprotein cholesterol (HDL-C), urine creatinine (UCr), uric acid (UA), serum creatinine (Scr), urea albumin creatinine ratio (UACR), blood urea nitrogen (BUN), and hypertension. Hypertension is characterized by a self-reported history of hypertension, the utilization of antihypertensive medications, or by achieving a mean systolic blood pressure of 140 mmHg or greater and/or a mean diastolic blood pressure of 90 mmHg or greater.

### 2.5 Statistical analysis

Continuous variables were reported as mean ± standard deviation (SD), while categorical variables are expressed as frequency (%). One-way analysis of variance (ANOVA) was utilized for the analysis of continuous variables, while the chi-square test was applied to categorical variables. We performed univariate and multivariate analyses to investigate the relationship between BRI and kidney stones. When considering BRI as a continuous variable, we utilized a linear regression model for the computation of the odds ratio (OR); correspondingly, logistic regression models were applied for categorical variables. Model 1 was not adjusted. Model 2 only adjusted for Age and Gender. Model 3 adjusted for Age, Gender, BMI, Weight, Education level, Smoking status, Marital status, HDL-C, Vigorous work activity, Moderate work activity, Physical Activity-Walk or bicycle, Vigorous recreational activities, Moderate recreational activities, Minutes sedentary activity, UCr, Scr, UA, UACR, BUN, TC, and Hypertension. To achieve statistical significance, a P value of < 0.05 was required. All estimates were calculated considering the NHANES weights. Weighting was applied to all analyses in accordance with the NHANES analysis guidelines. In this study, all statistical analyses were performed using R version 4.2.3 (R Foundation for Statistical Computing, Vienna, Austria) and SPSS version 23.0 (SPSS Inc., Chicago, IL, USA).

## 3. Results

### 3.1 Characteristics of the study population

Table 1 shows the results of comparing levels of the BRI and key demographic variables between diabetic patients with and without kidney stones. Among all participants (n = 3558), the mean BRI in the group with kidney stones was 7.18, which was higher than that in the group without kidney stones (7.18 vs. 6.95, p < 0.05). Whether diabetic patients have kidney stones or not is associated with various factors, including age, gender, weight, BMI, BRI, HDL-C, TC, UCr, Scr, UACR, BUN, marital status, engagement in strenuous work activities, cycling or bicycle use, and smoking status (p < 0.05). Diabetic patients with kidney stones were generally older and heavier and had higher BMI, lower HDL-C and TC levels, and elevated UCr, Scr, UACR, and BUN compared to those without kidney stones (p<0.05). Furthermore, diabetic patients with kidney stones were more likely to be male, possess an education level of at least some college or an associate degree, and be married or cohabiting.

**Tabel 1.**
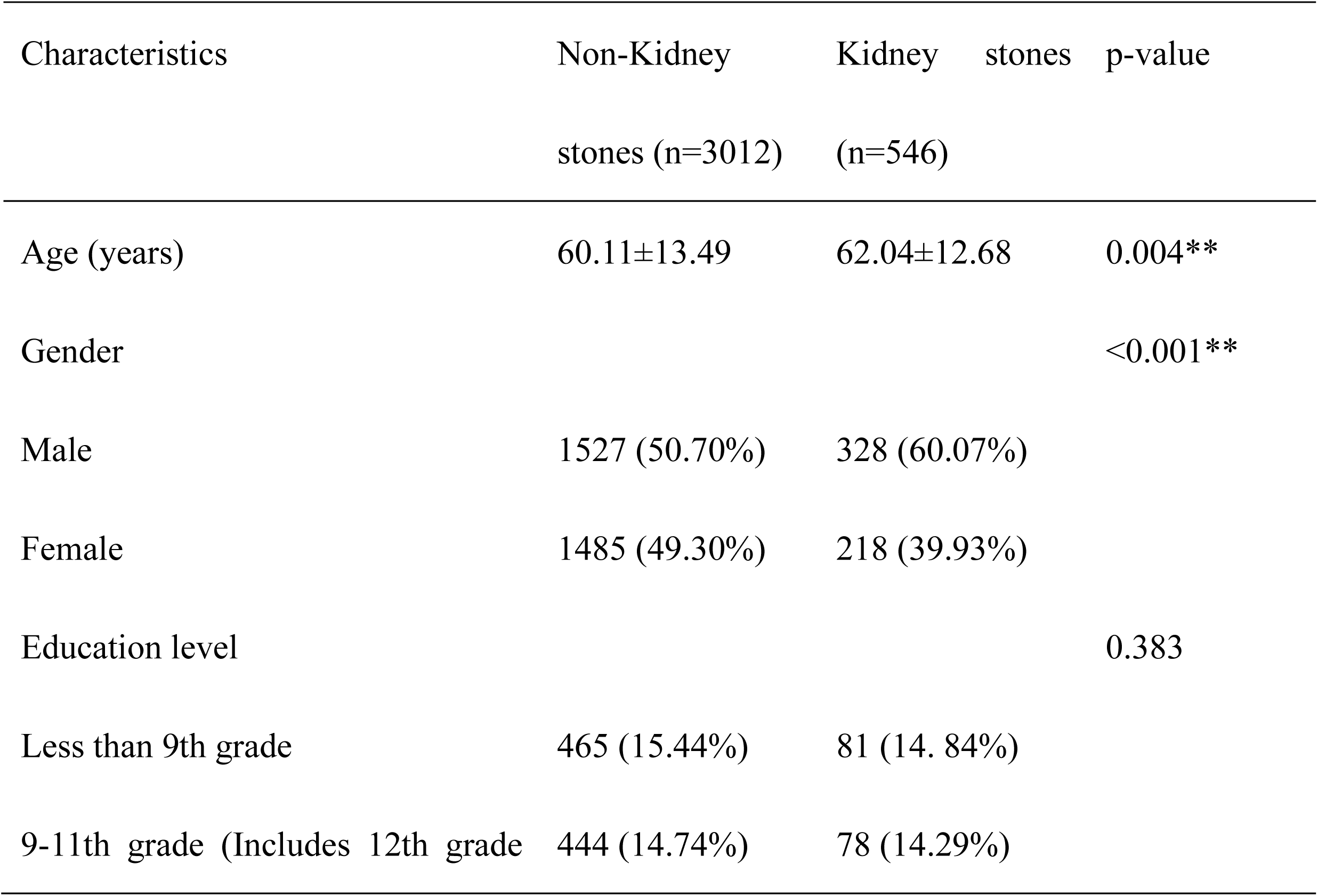
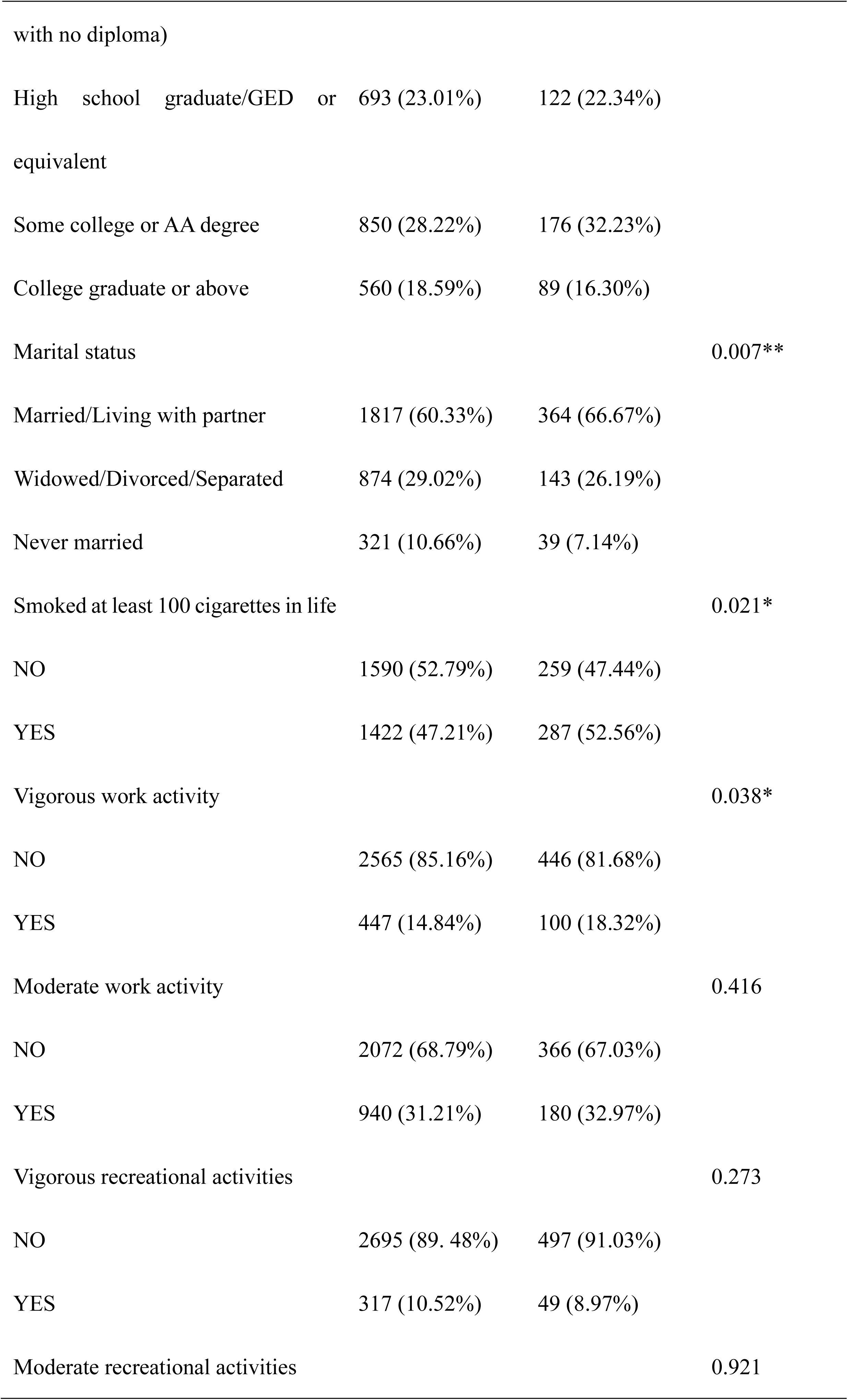
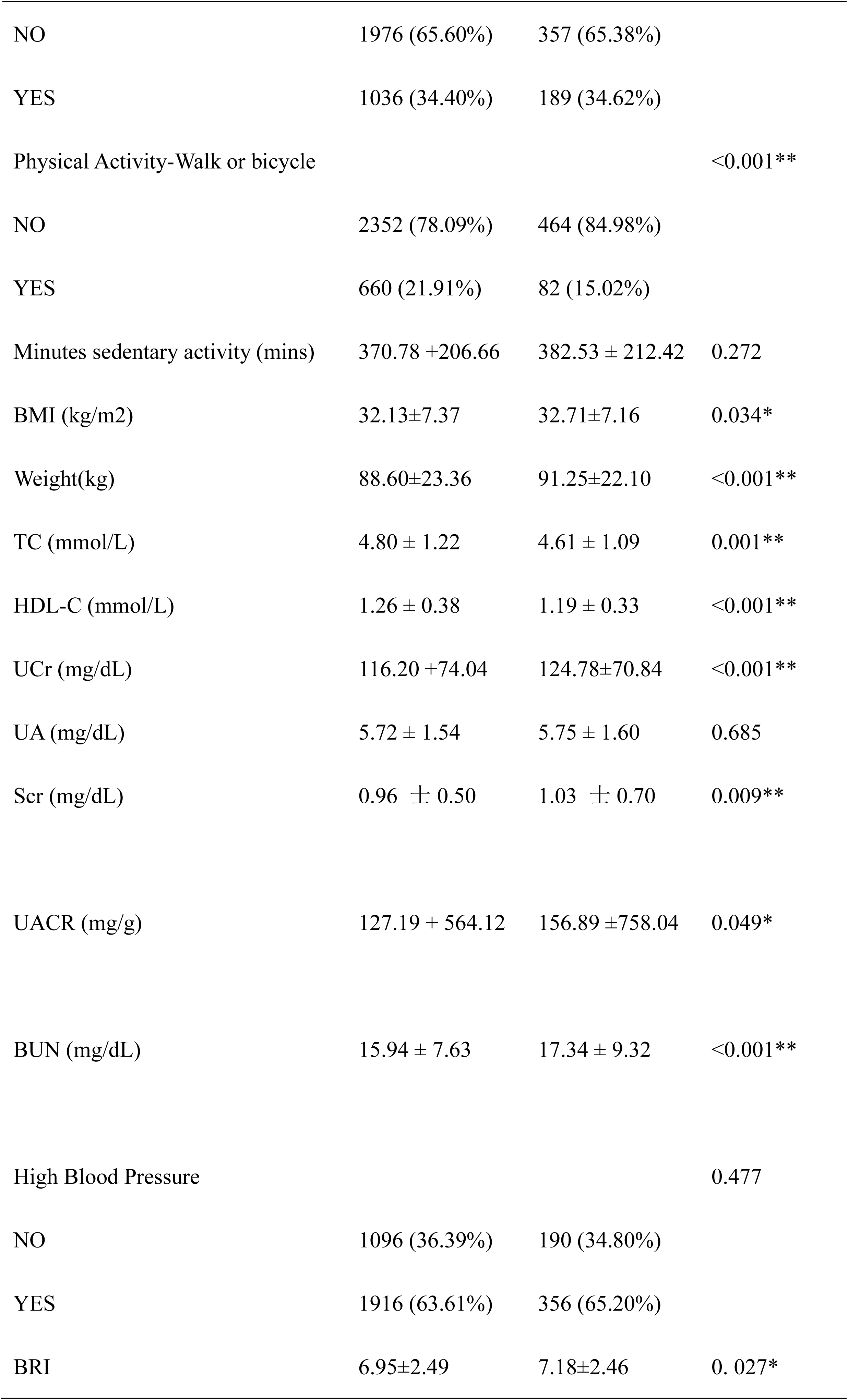
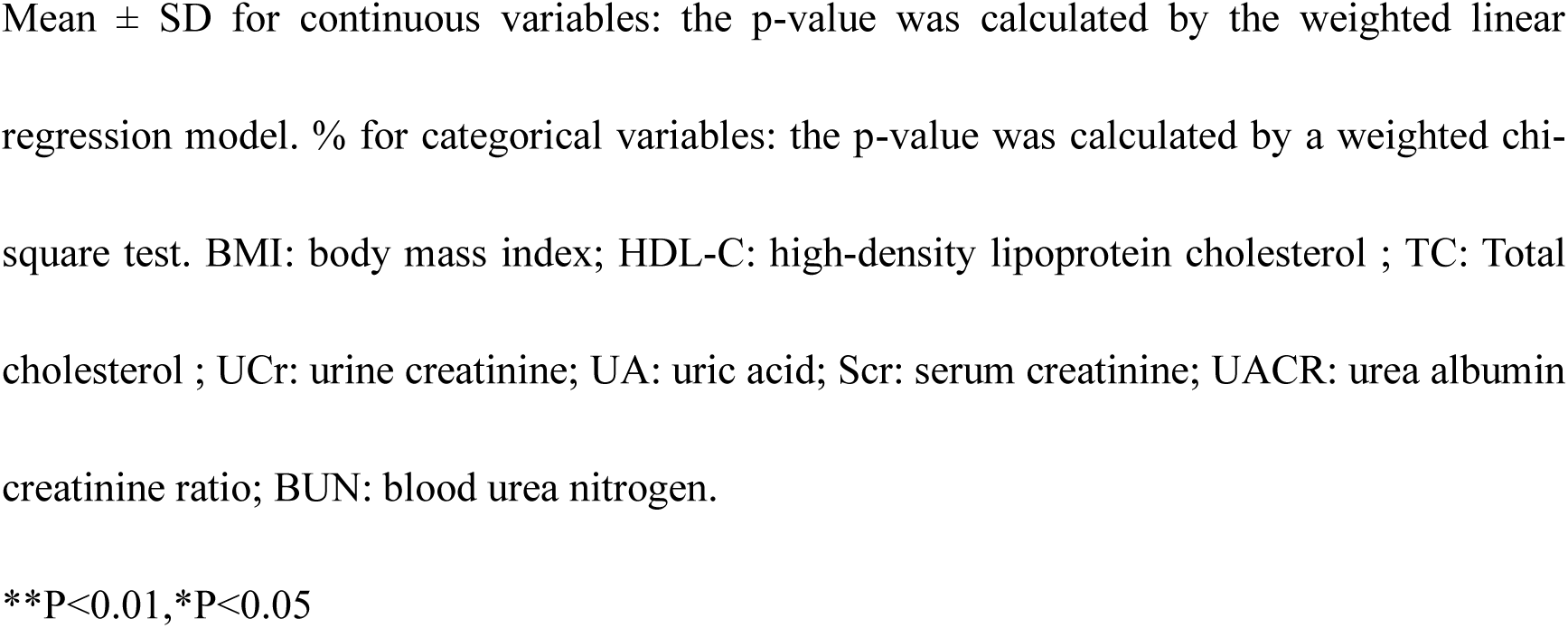
Baseline characteristics of the study participants.

### 3.2 Association between BRI and Kidney stones

Table 2 presents the results of the three logistic regression models. The results showed that for each unit increase in BRI among diabetic patients, the risk of kidney stones grew by 4%(OR=1.04, 95% CI: 1.00-1.07), 7% (OR=1.07, 95% CI: 1.03-1.11, P<0.001), and 5% (OR=1.05, 95% CI: 1.00-1.09; P<0.05), respectively. When BRI was divided into quartiles, this association remained robust. In Model 2, the prevalence in Q4 was significantly higher than in Q1 by 50% (OR=1.50, 95% CI: 1.14-1.98; P<0.05). At the same time, compared to Q1, the incidence of kidney stones in the three model groups in Q3 increased by 41%(OR=1.41, 95% CI: 1.08-1.83; P<0.05), 52%(OR=1.52, 95% CI: 1.16-1.99; P<0.05), and 37%(OR=1.37, 95% CI: 1.03-1.81; P<0.05), respectively.

**Table 2.**
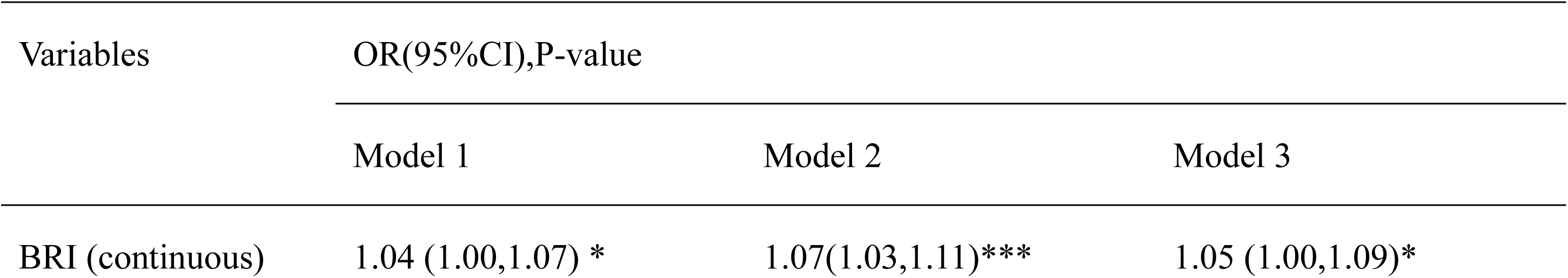
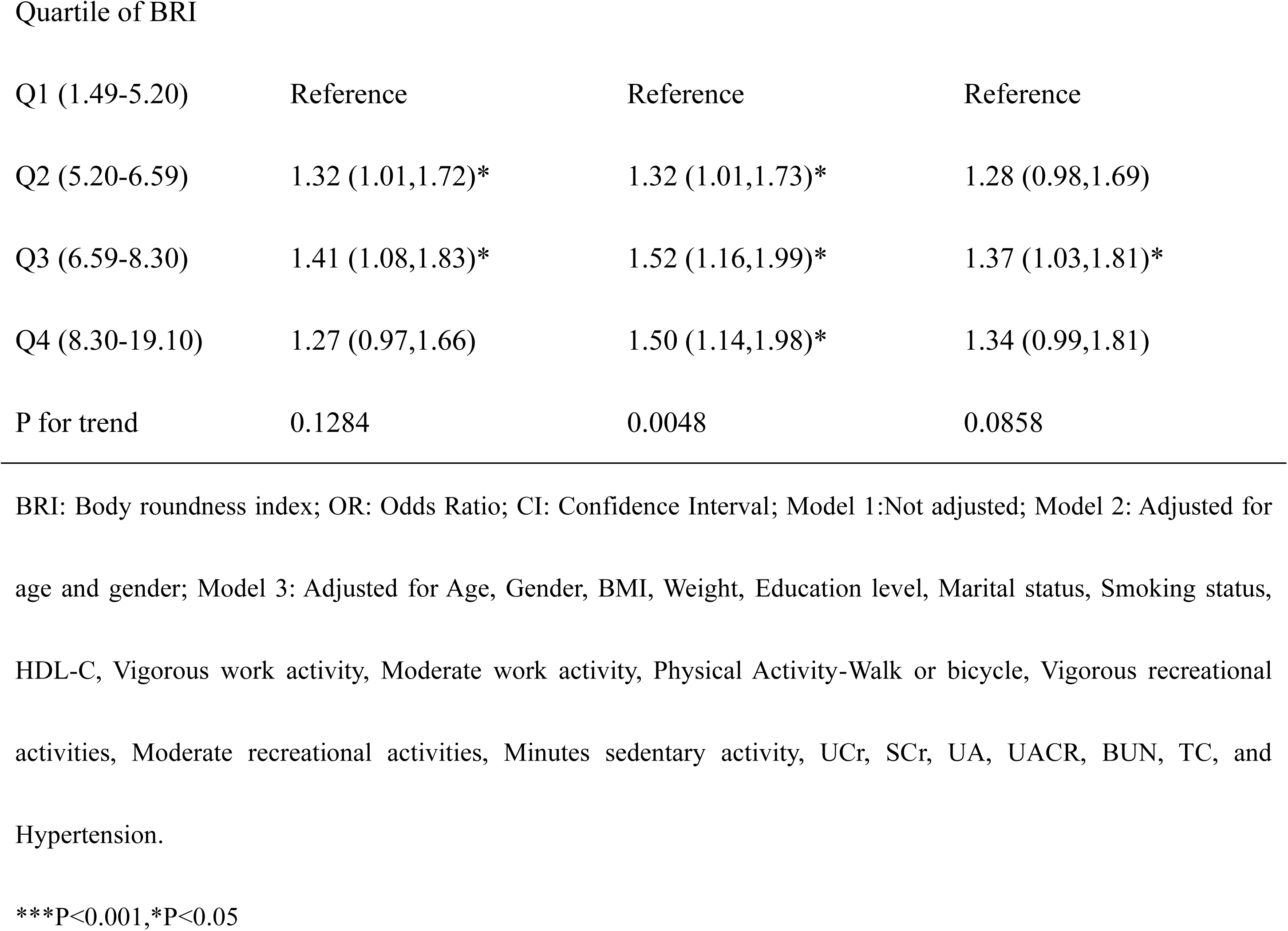
The association between BRI and Kidney stones.

| Variables | OR(95%CI),P-value |  |  |
| --- | --- | --- | --- |
|  | Model 1 | Model 2 | Model 3 |
| BRI (continuous) | 1.04 (1.00,1.07) * | 1.07(1.03,1.11)*** | 1.05 (1.00,1.09)* |

Quartile of BRI
| Q1 (1.49-5.20) | Reference | Reference | Reference |
| --- | --- | --- | --- |
| Q2 (5.20-6.59) | 1.32 (1.01,1.72)* | 1.32 (1.01,1.73)* | 1.28 (0.98,1.69) |
| Q3 (6.59-8.30) | 1.41 (1.08,1.83)* | 1.52 (1.16,1.99)* | 1.37 (1.03,1.81)* |
| Q4 (8.30-19.10) | 1.27 (0.97,1.66) | 1.50 (1.14,1.98)* | 1.34 (0.99,1.81) |
| P for trend | 0.1284 | 0.0048 | 0.0858 |

BRI: Body roundness index; OR: Odds Ratio; CI: Confidence Interval; Model 1: Not adjusted; Model 2: Adjusted for age and gender; Model 3: Adjusted for Age, Gender, BMI, Weight, Education level, Marital status, Smoking status, HDL-C, Vigorous work activity, Moderate work activity, Physical Activity-Walk or bicycle, Vigorous recreational activities, Moderate recreational activities, Minutes sedentary activity, UCr, SCr, UA, UACR, BUN, TC, and Hypertension.
\*\*\*P<0.001, \*P<0.05

### 3.3 Subgroup analysis

To further elucidate the stability of the results, subgroup analyses were performed. Interaction tests showed that the p for interaction>0.05 in the following variables: weight, BMI, Gender, age, HDL-C, Vigorous work activity, Moderate work activity, Physical Activity-Walk or bicycle, Vigorous recreational activities, Moderate recreational activities, Minutes sedentary activity, Smoked at least 100 cigarettes in life, TC, UCr, UA, Scr, UACR, BUN, Hypertension, education level and Marital status. This showed that these variables did not influence the relationship between BRI and kidney stones in diabetic patients, thus confirming the stability of the relationship between BRI and the prevalence of kidney stones in diabetic patients (Table 3).

**Tabel 3.**
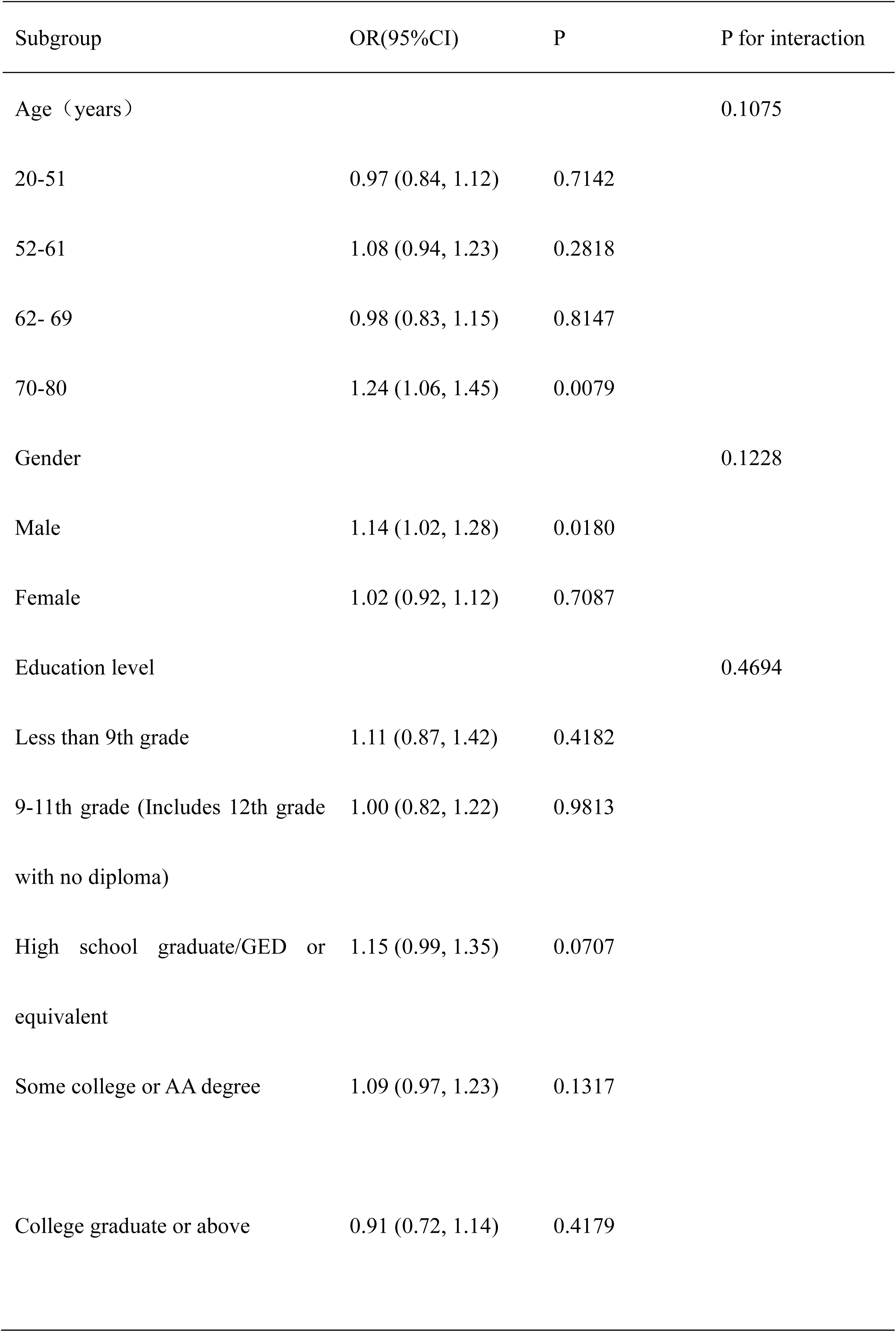
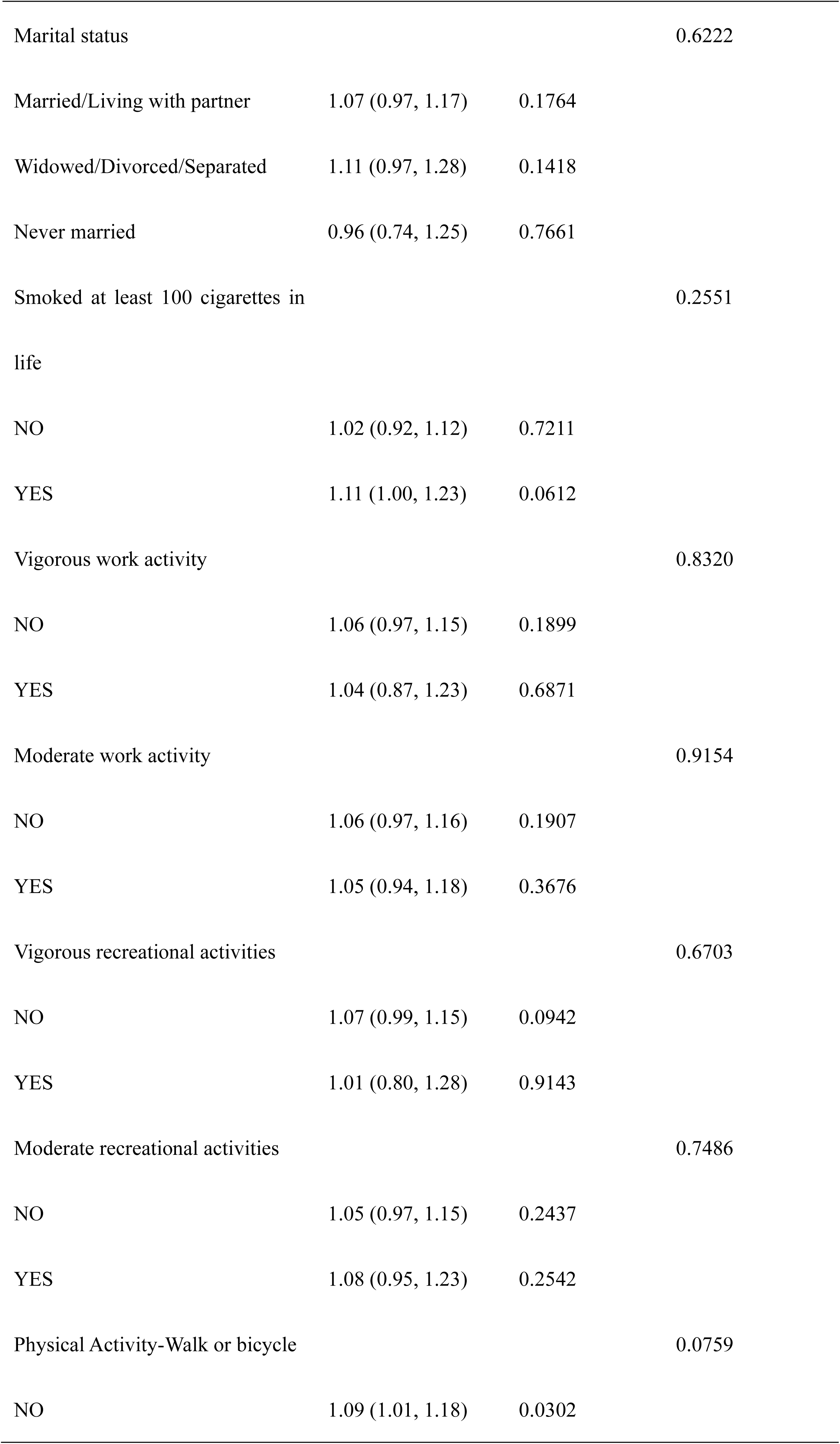
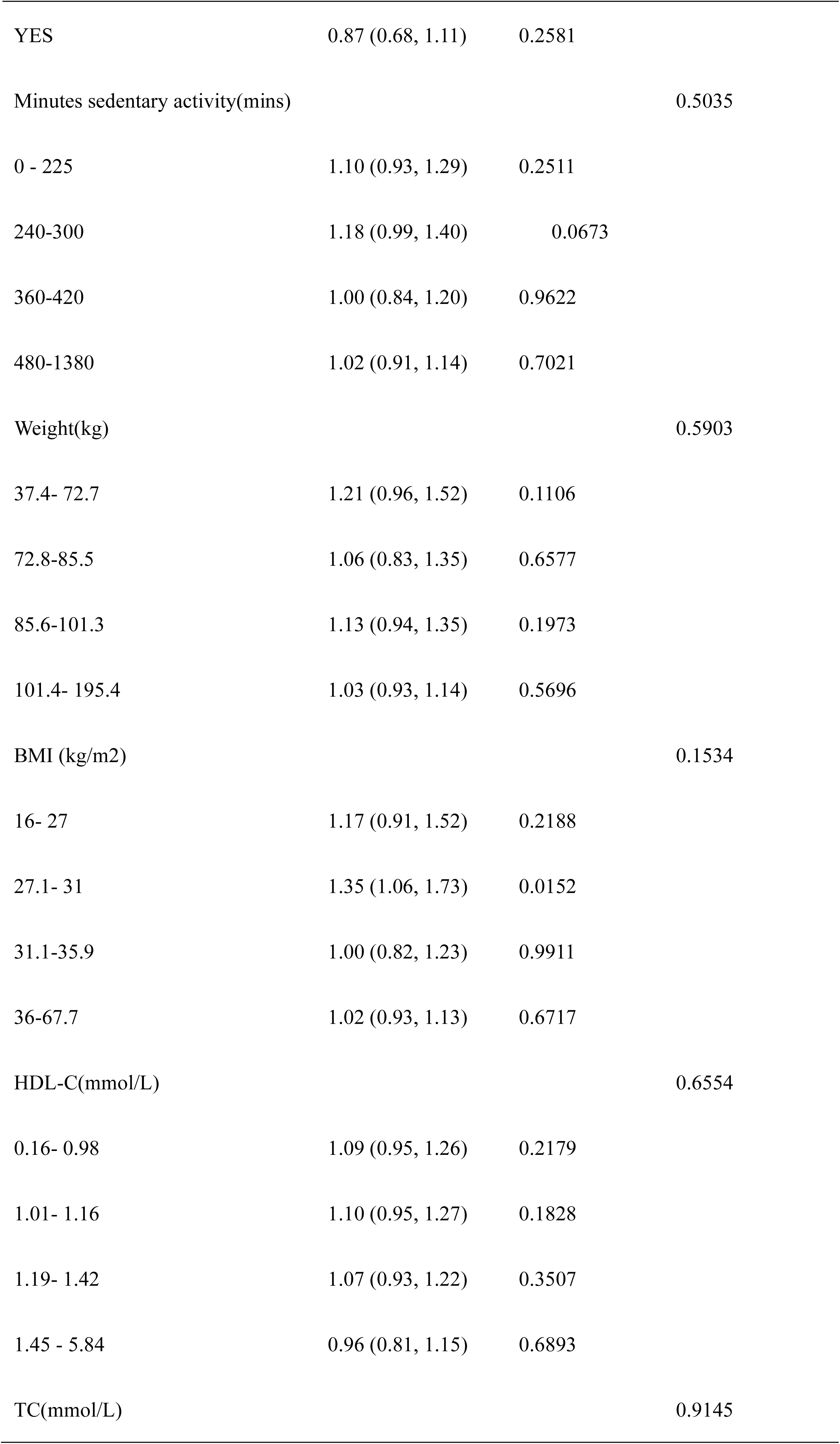
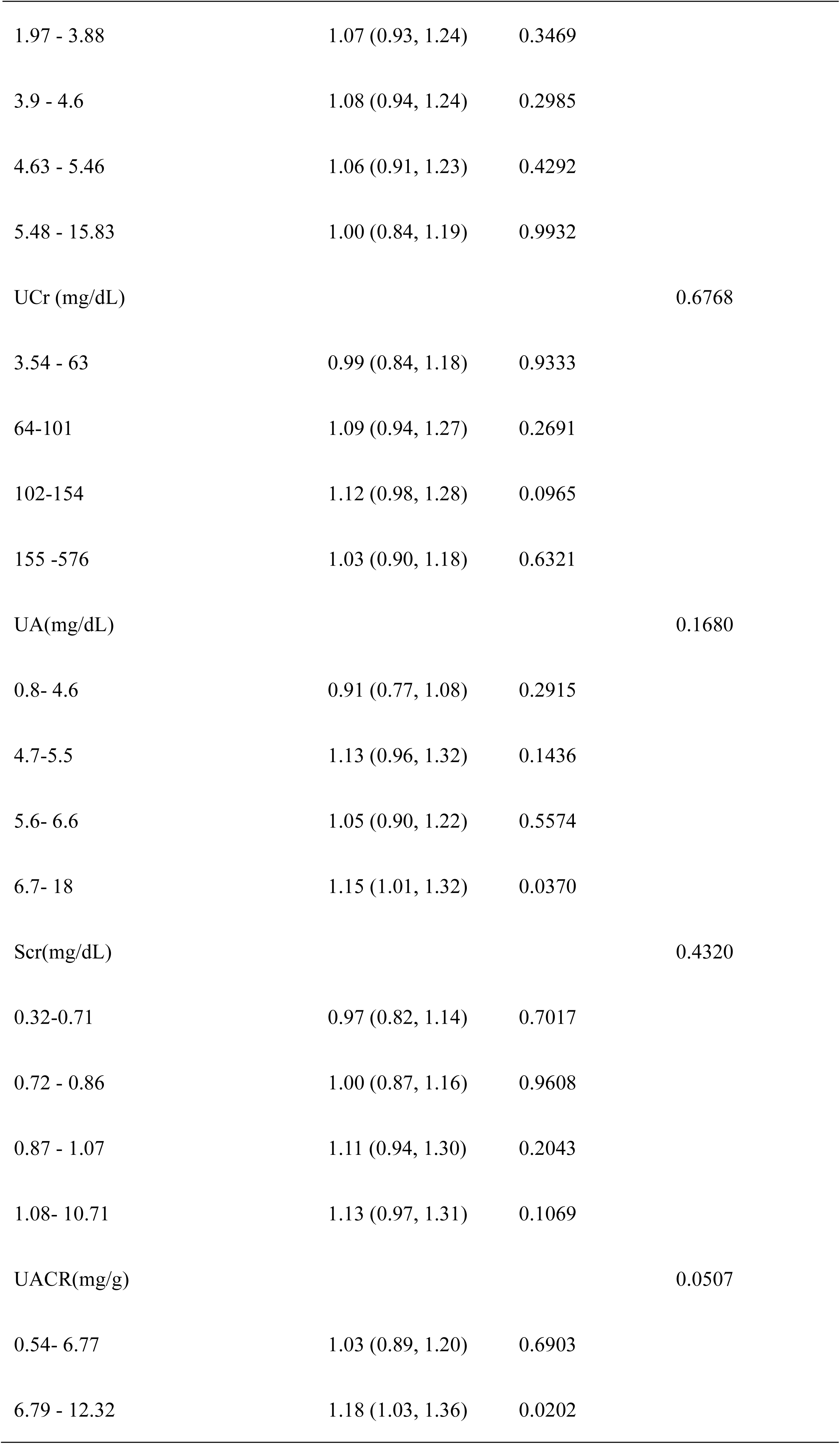
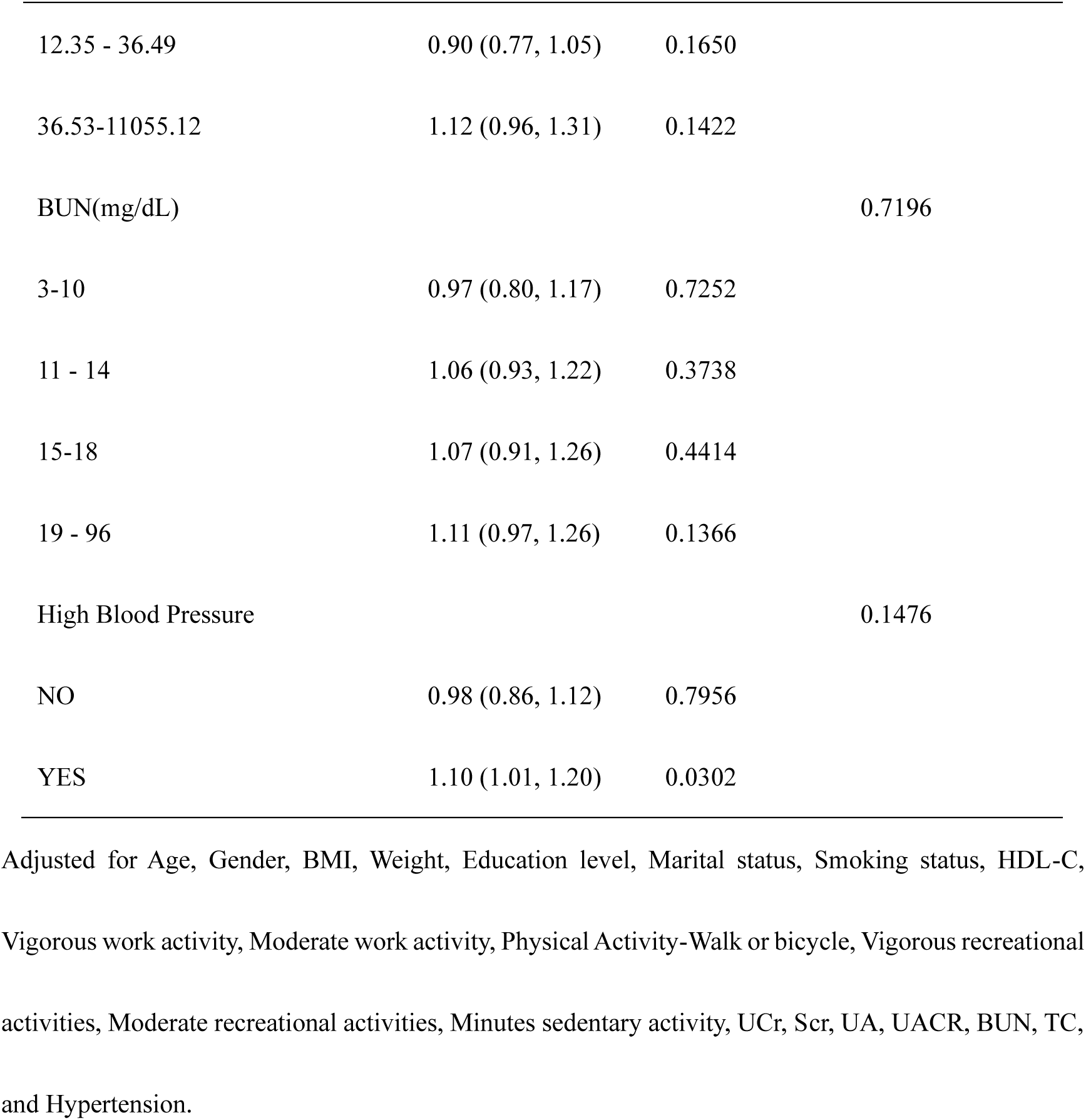
Subgroup analysis of the association between BRI and Kidney stones.

## 4. Discussion

This study represents the pioneering attempt to explore the link between BRI and the occurrence of kidney stones in diabetic patients. We found that the BRI of diabetic individuals with kidney stones was higher than that of those without kidney stones, and the data revealed a strong and a significantly positive association between BRI and kidney stone prevalence. This relationship remained robust in subgroup analyses.

Currently, there are no articles specifically investigating the correlation between BRI and the prevalence of kidney stones in diabetic populations, but a study by Xudong Hu et al. (10)indicated that the average BRI level in kidney stone patients (Mean ± SD = 6.11 ± 0.06) was substantially higher than that in non-kidney stone individuals (Mean ± SD = 5.36 ± 0.03). Gaoteng Lin et al.(15)also reached the same conclusion, noting that an increase in BRI values leads to a higher risk of developing kidney stones in men aged 20-39 years (OR = 1.34, 95% CI 1.06-1.69) and 40-59 years (OR = 1.29, 95% CI 1.09-1.53). Similarly, in women aged 40- 59 years, the prevalence of kidney stones increased with rising BRI values (OR = 1.23, 95% CI 1.07-1.41). Compared to BMI, BRI is more accurate in identifying the risk of kidney stones(10).

Research by Rahman IA et al. (16) indicates that there is a certain association between diabetes and an increased likelihood of developing kidney stones, making it particularly important to identify and manage high-risk populations with diabetes and kidney stones early in clinical practice. Our study shows that in diabetic patients, higher BRI levels correlate with a heightened risk of developing kidney stones (P < 0.05), suggesting that BRI in diabetic patients may serve as a predictive indicator for assessing the risk of kidney stones in this population. However, the mechanisms underlying the relationship between elevated BRI and the occurrence of kidney stones in diabetic patients remain unclear. We speculate that this may be due to the metabolic disorders often associated with diabetes, including hyperglycemia, dyslipidemia, and insulin resistance, which could all affect kidney function and consequently increase the risk of kidney stones. More importantly, studies have shown that obesity, a risk factor for kidney stones, increases the likelihood of developing these stones(17,18). The prevalence of obesity is often higher among individuals with diabetes, primarily due to insulin resistance. In response to insulin resistance, the pancreas increases insulin secretion, leading to hyperinsulinemia. This surge in insulin levels can stimulate fat deposition, especially in the abdominal area, thereby exacerbating obesity and increasing the incidence of kidney stones. Therefore, we believe that obesity is one of the manifestations of glucose and lipid metabolism disorders in diabetic patients and is also a risk factor for the occurrence of kidney stones. If an indicator could be identified to assess the obesity levels in diabetic patients, it would aid in better understanding and preventing the occurrence of kidney stones in diabetes, thereby offering new strategies to reduce the risk of kidney stones in this vulnerable population.

Currently, the indicators for assessing visceral fat in the body are primarily BMI and WC, but they provide limited information on fat distribution(19,20). In contrast, BRI can comprehensively assess the percentage of visceral fat and total body fat, thus providing a more comprehensive picture of fat distribution in diabetic patients. Therefore, using BRI instead of BMI and WC may provide more accurate information for assessing fat distribution in diabetic patients. Previous studies have shown that BRI levels in diabetic and prediabetic populations are higher than those in non-diabetic populations(21,22). This may be due to the fact that in diabetic patients, hyperglycemia and insulin resistance trigger increased fat synthesis in the liver and accelerated fatty acid metabolism in adipose tissue, which ultimately leads to abnormal lipid levels(23,24). We propose that if lifestyle interventions and pharmacological treatments fail to to control lipid metabolism disorders in diabetic patients effectively, it might instigate stress on the endoplasmic reticulum along with oxidative stress reactions. These responses could lead to decreased gene expression and secretion, as well as increased apoptosis of pancreatic β-cells, ultimately impairing β-cell function. As a result, impaired β-cell function reduces insulin sensitivity and secretion, contributing to persistent hyperglycemia and accelerating the development of kidney stones(25).

Lipocalin, as a protective adipokine, is influenced by visceral fat levels. When visceral fat levels increase, adiponectin concentrations decrease, while levels of pro-inflammatory cytokines (such as tumor necrosis factor and interleukin-6) rise, further exacerbating insulin resistance(26). Similarly, a meta-analysis by Geraghty R et al. that included cohort studies, cross-sectional studies, and case-control studies indicated that the incidence of kidney stones is significantly increased in patients with diabetes. This occurs as insulin resistance diminishes both the production and the transport of renal ammonium in the proximal renal tubules while increasing the reabsorption of urinary bicarbonate and sodium, leading to excessive urine acidification and a decrease in urine pH(27). The increase in fatty acids further lowers urine pH. The highly acidic urine produced by obese patients reduces the saturation of uric acid and calcium oxalate complexes in the urine, resulting in the creation of uric acid stones. Additionally, insulin resistance decreases urinary citrate concentration, which not only decreases urine pH and encourages the development of uric acid stones but also aids in the formation of calcium oxalate stones. Once uric acid crystals form, urinary oxalate and calcium ions can also promote the creation of calcium oxalate stones via a process of heterogeneous nucleation(28,29). In terms of lipids, our findings suggested that diabetic patients with kidney stones had lower HDL-C levels (Mean±SD=1.19±0.33) than those without kidney stones (Mean±SD=1.26±0.38) (p<0.001). Besiroglu et al. analyzed 11 studies on the correlation between urinary stones and dyslipidemia and found that lower HDL-C levels significantly increased the risk of urinary stones(30). Data from a study involving 38,617 U.S. adults indicated that HDL-C levels were significantly lower in individuals with kidney stones compared to those without, and that lower HDL-C levels were a risk factor for kidney stone formation among U.S. adults, while TC, triglycerides (TG), and LDL-C showed no association with stone formation(31). These findings are consistent with our research. This could be due to dyslipidemia’s contribution to chronic inflammation and oxidative stress, factors that are instrumental in the development of urinary stones. Previous studies have shown that inflammation resulting from metabolic changes promotes crystal formation in the kidneys of obese mice with metabolic syndrome(32). Patients with diabetes exhibit an increased prevalence of kidney stones relative to the general population, likely due to insulin resistance and dyslipidemia. Currently, there are no established predictors of kidney stone risk specifically for the diabetic population. If an indicator can be used to predict the risk of kidney stones in diabetes mellitus. In that case, appropriate clinical strategies can be developed to effectively intervene in patients and decrease the occurrence of kidney stones in the diabetic individuals. Our research demonstrated that the prevalence of kidney stones among people with diabetes increased in parallel with rising BRI levels, suggesting that BRI could serve as a valuable predictor of kidney stone risk in this population.

The advantages of this study are as follows: 1. This study is based on NHANES data from 2011 to 2018, with a large sample size that enhances the representativeness and statistical power of the findings. 2. This study comprehensively considers data, including demographic characteristics, lifestyle habits, clinical measurements, and biochemical indicators, effectively controlling for various potential confounding factors. 3. As the first study to explore the association between BRI and kidney stones in the diabetic population, it reveals that metabolic disturbances in this population may exacerbate the risk of kidney stones, making the findings particularly significant for the prevention and treatment of kidney stones in individuals with diabetes. The limitations of this study are as follows: 1. Due to the cross-sectional nature of NHANES data, causality cannot be established, and only the association between BRI and kidney stones can be observed. 2. Some information in the study, such as the diagnosis of diabetes and lifestyle factors, relies on self-reporting by participants, which may introduce recall bias. 3. Individuals with incomplete data were excluded from the analysis, which may affect the selectivity of the sample and the generalizability of the results. 4. Since NHANES data primarily comes from a non-institutionalized population in the United States, the findings may be limited by racial and regional characteristics, restricting their applicability on a global scale. Future research needs to validate these findings in different populations and consider more potential confounding factors to enhance the broader applicability of the results.

## 5. Conclusion

Utilizing NHANES data from 2011 to 2018, this study is the first to explore the relationship between BRI and kidney stones in a diabetic population, providing new insights. This study found that BRI was higher in diabetic patients with kidney stones compared to those without. Furthermore, a significant positive correlation was observed between BRI and the prevalence of kidney stones in the diabetic population, suggesting that BRI may serve as a valid indicator for assessing the risk of kidney stones in this group and highlighting the important role of visceral adiposity in kidney stone pathogenesis within the context of diabetes-related metabolic disorders.

## Data Availability

All relevant data are within the manuscript and its Supporting Information files.

## Author contributions

**Conceptualization:** Chongsong Cui.

**Data curation:** Chongsong Cui, You Peng, Ying Zhao.

**Formal analysis:** Chongsong Cui.

**Investigation:** Feiin Chan, Qiqi Ren.

**Methodology:** Chongsong Cui.

**Project administration:** Chongsong Cui.

**Resources:** Feiin Chan.

**Supervision:** Zhenjie Liu.

**Validation:** Feiin Chan.

**Visualization:** Qiqi Ren.

**Writing– original draft:** Chongsong Cui, You Peng, Ying Zhao.

**Writing– review & editing:** Zhenjie Liu

## Consent for publication

Not applicable

## Ethical statement

The studies involving human participants were reviewed and approved by NCHS Ethics Review Board. The patients/participants provided their written informed consent to participate in this study.

## Acknowledgments

We are grateful for the National Health and Nutritional Evaluation Survey being made accessible to the entire population by the National Center for Medical Research at the Institute of Prevention and Control of Disorders.

## Data availability

The survey data are publicly available on the internet for data users and researchers throughout the world ( www.cdc.gov/nchs/nhanes/ ).

